# Reduction in time delay of isolation in COVID-19 cases in South Korea

**DOI:** 10.1101/2020.04.03.20051847

**Authors:** Sukhyun Ryu, Cheolsun Jang, Baekjin Kim

## Abstract

Korean public health authorities raised the public alert to its highest level on February 23, 2020 to mitigate the 2019 novel coronavirus disease epidemic. We have identified that the mean delay from symptom onset to isolation was reduced to one day after raising the alert. Vigilance can reduce this interval.

---

Since the first case of the 2019 novel coronavirus disease (COVID-19) was identified in South Korea on January 20, 2020 [1], COVID-19 has infected 9,887 people and killed 165 in South Korea as of April 1, 2020 [2]. In the early phase of the COVID-19 epidemic in Korea, Korean public health authorities mainly conducted contact tracing of confirmed cases and quarantining of confirmed and suspected cases [1]. However, as the number of COVID-19 cases increased, Korean public health authorities raised the infectious disease alert to red alert on February 23, 2020 [3]. Central disaster and safety countermeasures were set up and addressed the public alert to report illness related to COVID-19 to public health authorities for screening.

To assess the response’s impact during the highest alert phase in Korea, we obtained data on 576 confirmed COVID-19 cases reported from the Korea Centers for Disease Control and Prevention, the city department of public health, and news reports for February 1–March 30, 2020. We extracted information about confirmed cases using a structured data-extraction form. We used case-based data on the date of exposure of confirmed cases, date of illness onset, and date of isolation. We compared the time interval from illness onset to isolation before and during the red alert (after February 23, 2020), assuming that the number of intervals was log-normal, Weibull, or gamma-distributed (Appendix). Further, we conducted a similar analysis to identify the interval from exposure to isolation. The range of intervals was reported with 95% confidence intervals (2.5th and 97.5th percentiles). To identify the difference between these intervals in the early epidemic and during the red alert, we conducted a Mann–Whitney U test. All statistical analyses were performed in R version 3.0.2 (R Foundation for Statistical Computing).

Of the 576 laboratory-confirmed cases reported (6% of the 9,786 Korean COVID-19 cases as of March 31, 2020), 211 included the date of exposure, illness onset, or isolation and were analyzed (Appendix). The mean delay between symptom onset and isolation was 4.3 days (95% CI 0.5–11.3 days) in the early phase and 3.3 days (95% CI: 0.5–9.4 days) during the red alert. This reduction was significantly lower than in the early phase of COVID-19 epidemics in Korea (*p*-value = 0.02). The mean delay from exposure to a confirmed case to isolation was 7.2 days (95% CI 1.7–14.4 days) before the red alert and 6.5 days (95% 1.7–15.7 days) during the red alert (Figure). Therefore, the mean delay was shortened by 0.8 days; however, this was not significantly lower (*p*-value = 0.07).

**Figure.**
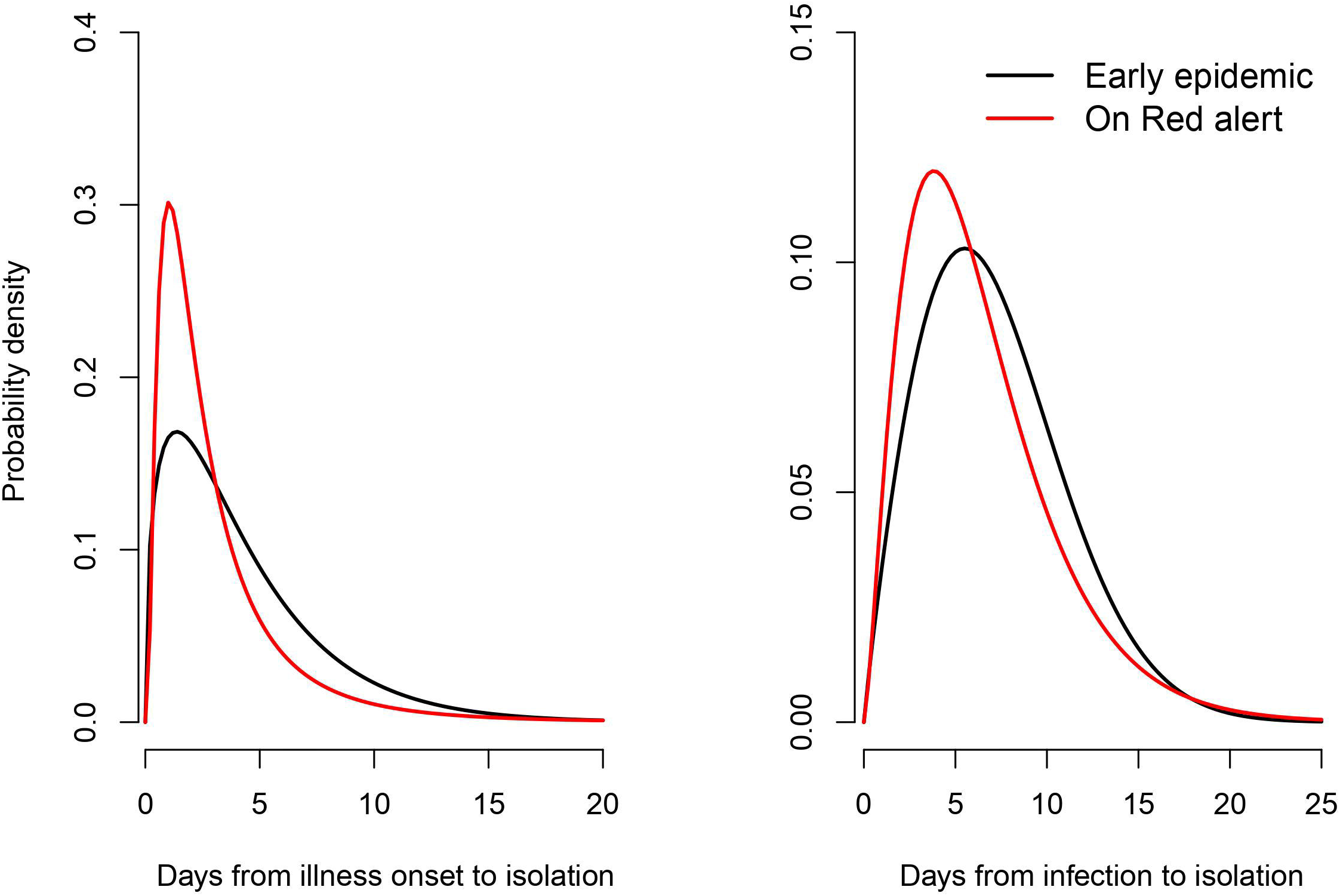
Distribution of interval from symptom onset to isolation and exposure to isolation. (A) Symptom onset to isolation data from 211 cases in South Korea early in the epidemic. The black line is the estimate before the red alert was implemented on February 23, 2020 and indicates a fitted gamma distribution. The red line is the estimate during the red alert and indicates a fitted log-normal distribution. (B) Data of exposure to isolation from 174 cases in South Korea. The black line is the estimate before the red alert was implemented on February 23, 2020 and indicates a fitted gamma distribution. The red line is the estimate during the red alert and indicates a fitted log-normal distribution.

Most COVID-19 transmissions occur during the presentation of symptoms of cases; therefore, we believe that COVID-19-affected countries are making efforts to reduce the time delay from symptom onset to isolation by strengthening countrywide surveillance and control programs for COVID-19. In Korea, to enhance the surveillance, public health authorities have operated 80 drive-through screening centers (operated since February 23, 2020) and have designated 341 private hospitals as public relief hospitals for COVID-19 (operated since February 25, 2020) [4, 5]. This provided easier access to screening tests for suspected cases of COVID-19 in the community compared to the early phase and could reduce the interval from symptom onset to isolation.

The delay from illness onset to isolation early in the epidemic in Korea is relatively shorter than that in China (mean estimate of 5.8 days, 95% CI 4.3–7.5 in early January 2020) [6]. However, more efforts are needed from public health authorities and the public to prevent the potential spread of COVID-19 in communities since the infectiousness of COVID-19 begins before illness onset [7].

Our study has some limitations. We estimated the time delay based on self-reported data and obtained data from various sources. Additional studies should incorporate more detailed information from additional laboratory-confirmed cases.

Efforts to reduce this interval are ongoing in South Korea and vigilance in other countries can reduce this interval.

## Data Availability

We obtained epidemiological data from publicly available data sources (news articles and press releases)

## Acknowledgments

We appreciate the Korean public health authorities’ response to COVID-19 in Korea.

## About the Author

Dr. Ryu is an assistant professor of preventive medicine at Konyang University, Daejeon, South Korea. His research interests include infectious disease epidemiology with focus on influenza and public health interventions.

